# Stroke Probability Prediction from Medical Survey Data: AI-Driven Analysis with Insightful Feature Importance using Explainable AI (XAI)

**DOI:** 10.1101/2023.11.17.23298646

**Authors:** Simon Bin Akter, Sumya Akter, Tanmoy Sarkar Pias

## Abstract

Prioritizing dataset dependability, model performance, and interoperability is a compelling demand for improving stroke risk prediction from medical surveys using AI in healthcare. These collective efforts are required to enhance the field of stroke risk assessment and demonstrate the transformational potential of AI in healthcare. This novel study leverages the CDC’s recently published 2022 BRFSS dataset to explore AI-based stroke risk prediction. Numerous substantial and notable contributions have been established from this study. To start with, the dataset’s dependability is improved through a unique RF-based imputation technique that overcomes the challenges of missing data. In order to identify the most promising models, six different AI models are meticulously evaluated including DT, RF, GNB, RusBoost, AdaBoost, and CNN. The study combines top-performing models such as GNB, RF, and RusBoost using fusion approaches such as soft voting, hard voting, and stacking to demonstrate the combined prediction performance. The stacking model demonstrated superior performance, achieving an F1 score of 88%. The work also employs Explainable AI (XAI) approaches to highlight the subtle contributions of important dataset features, improving model interpretability. The comprehensive approach to stroke risk prediction employed in this study enhanced dataset reliability, model performance, and interpretability, demonstrating AI’s fundamental impact in healthcare.

## I Introduction

Stroke, a significant cause of death and disability world-wide, can benefit from AI’s capacity to quickly analyze massive details, detect complicated patterns, and improve preventive strategies [1]. Integrating artificial intelligence (AI) into stroke risk assessment optimizes healthcare resource allocation, lowering the burden of stroke-related morbidity and death while improving patient outcomes.

Research on stroke risk prediction makes heavy utilization of AI techniques like machine learning and neural networks as well as the inclusion of many risk variables, including genetic markers, to improve predictability [2] [3] [4]. However, difficulties with generalization and model improvement still exist. In order to improve the area, future research should focus on the creation of approachable methods and the integration of novel risk factors.

The possibilities of AI in stroke risk prediction are being unlocked by examining new datasets, according to recent literature reviews [5] [6]. Complexity is increased by the requirement for model comparisons to determine the best methods. It is imperative to address the frequent issue of missing data and class imbalance in survey datasets. To further emphasize the clinical applicability of predictive characteristics and improve the interpretability of stroke risk prediction models, explainable AI (XAI) techniques are required [7] [8].

The 2022 Behavioural Risk Factor Surveillance System (BRFSS) dataset from the Centres for Disease Control and Prevention is employed in this study. To effectively handle missing data and data imbalances, this study includes substantial preprocessing approaches. The effectiveness of several machine learning and deep learning models in predicting the risk of stroke is thoroughly compared in the study. Additionally, it uses an Explainable AI (XAI) framework to shed clarification regarding the influence of various characteristics and the contributions of these to stroke risk prediction.

The key contributions are summarized below.

1. In the initial exploration of the BRFSS 2022 dataset, various AI approaches, including model fusion techniques, were systematically assessed for predictive performance in determining stroke risk.
2. Exploratory data analysis (EDA) has been performed to identify and visualize different patterns and statistics from the dataset.
3. A Machine Learning-based imputation technique has been introduced to predict a vast amount of feature-specific missing values based on the existing insights from the clean data.
4. Explainable AI (XAI) techniques are employed to identify and explain the most relevant input parameters in stroke risk prediction elucidating the impact.

In conclusion, this study addresses the fundamental obstacle of stroke risk assessment by applying powerful Artificial Intelligence (AI) algorithms to survey data from a medical setting. This study aims to improve healthcare outcomes by actively contributing to the development of a resilient diagnostics approach, opening the way for the early identification and successful management of strokes.

## II. Literature review

Disease risk prediction, as well as health behavior and habit analysis based on survey data, serve as essential for improving healthcare. It allows for the early identification of those who are at a higher risk of developing certain diseases, allowing for focused preventative measures and personalized healthcare regimens. Furthermore, this study provides useful insights into the variables impacting health behaviors and habits, which may help healthcare providers personalize treatments for better lifestyles. Furthermore, identifying widespread health conditions within certain groups aids resource allocation and healthcare policy creation.

Connie et al. [10] performed a detailed study utilizing BRFSS data to address health disparities while employing COVID-19 components in heart disease and stroke research. Ryan [11] investigated the link between police homicides and cardiovascular events including hypertension, diabetes, heart attack, and stroke using BRFSS and government records. Early on, Yashvanth et al. [12] used machine learning algorithms on BRFSS data to predict diabetes, stroke, and hypertension, focusing on data quality and model selection. Chuan et al. [13] assessed stroke risk prediction models in a diverse population, emphasizing the importance of improving modeling methodologies and risk factor inclusion in order to overcome racial discrepancies in stroke prediction.

Madhab et al. [14] used machine learning approaches such as Random Forest to predict and categorize stroke diseases based on risk indicators. Using logistic regression modeling, Debora et al. [15] investigated the incidence of stroke risk variables across rural-urban areas and their relationship to neighborhood socioeconomic level. Connie et al. [16] used BRFSS data to suggest a plan for the American Heart Association’s Statistical Update that focused on socioeconomic determinants of health, unfavorable pregnancy outcomes, and worldwide cardiovascular disease burden. Using data from the Florida Department of Health’s BRFSS survey, Marufuzzaman et al. [17] calculated stroke prevalence and predictors among those with prediabetes and diabetes. Neal et al. [18] used BRFSS data to examine smoking cessation in stroke and cancer survivors in the United States, adjusting for demographics and smoking-related variables. Phoebe et al. [19] looked studied antihypertensive medication usage and lifestyle factors in hypertensive stroke survivors, focusing on rural-urban differences and changes over time. Tarang et al. [20] used BRFSS data to predict frequent marijuana use and identify significant predictors, with Random Forest outperforming all others. Debayan et al. [21] investigated the association between behavioral features and prevalent illnesses, including work-life balance, physical activity, diet, tobacco/alcohol intake, and stroke history, using machine learning on BRFSS data.

A thorough investigation of cardiovascular disease, stroke, and associated variables using AI approaches is highlighted in the reviewed literature. Addressing data imbalances, creating reliable methods for handling missing data, incorporating Explainable AI (XAI) techniques for deeper insights, and performing more thorough model comparisons are notable research gaps that could improve the precision and applicability of predictive models in this domain. These developments are essential for expanding our knowledge of these diseases and creating more potent preventative and intervention plans.

## III. Dataset

The Behavioural Risk Factor Surveillance System (BRFSS) 2022 dataset was used in this study, which is a comprehensive repository of health-related information collected from surveys accomplished across the United States. The analysis and prediction of stroke risk stand out as a major emphasis area in this dataset. A wide range of demographic, lifestyle, and health-related characteristics are used to develop stroke risk prediction models [11] [12] [14] [16].

The primary objective of this research is to predict the risk of stroke. The selected target variable for this study shows whether individuals have ever had a stroke diagnosis. Notably, the study exhibits a severe class imbalance issue as there are many more data points for healthy subjects than for individuals with stroke. Taking into account variables like gender, age ranges, and race/ethnicity, the target class has been completely represented in **Fig 1**. At first, stroke instances are unbalanced, with more females being afflicted. The incidence of stroke is also higher in the elderly than in the young. Accordingly, the data indicates that white Americans have the highest rate of stroke instances. This study provides significant details on the distribution of stroke cases by gender, age, and ethnicity.

**Fig. 1:**
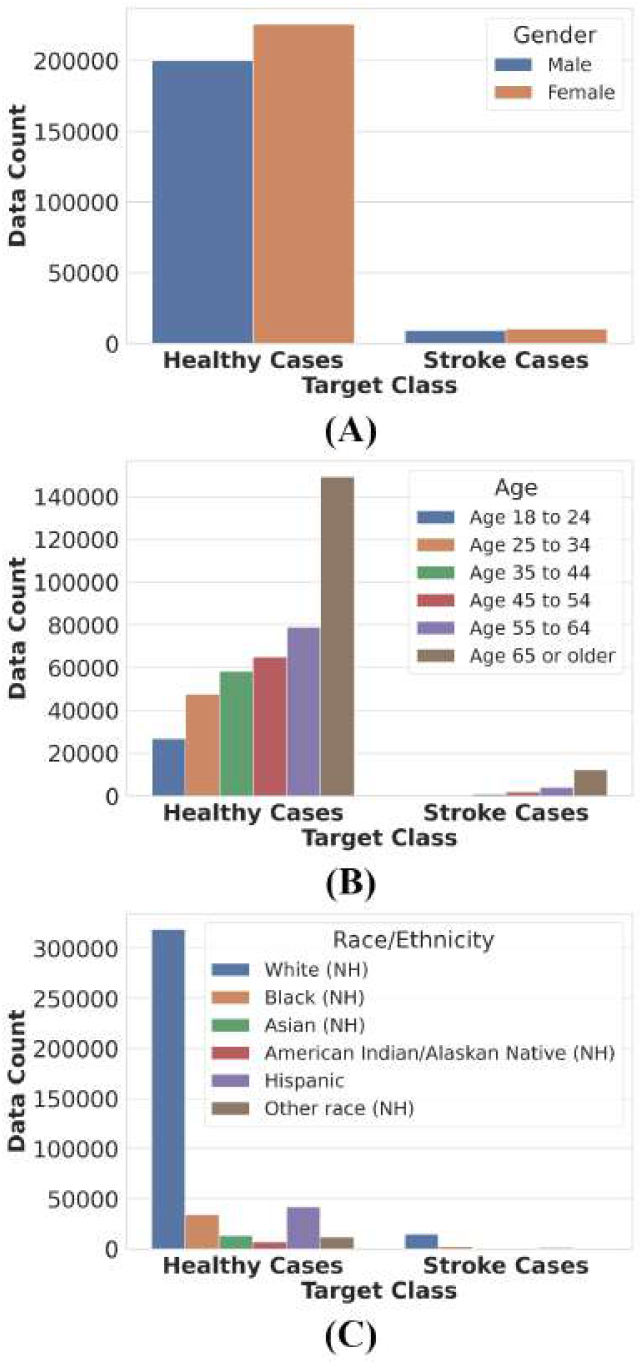
Representation of the target class by referring to different input characteristics including gender, age groups, and race or ethnicity. (A) The number of individuals classified by gender and health status is estimated in this section of the study. (B) The number of individuals classified by different age groups and health status is quantified in this area of the study. (C) The number of individuals classified by race or ethnicity and health status is tracked in this segment of the study. This section distinguishes between those who are healthy and those who have endured a stroke by pointing to different key input features.

## IV. Methodology

Predicting strokes employing medical survey data has significance as it allows for the early identification of patients who might be at high risk, allowing for prompt treatments and preventative actions [2] [4] [5] [6]. Stroke is a potentially fatal disorder that frequently results in significant disability and healthcare costs, yet many risk factors are controllable if identified early. By analyzing medical survey records, it can be possible to come up with focused policies for education, awareness, and healthcare resource allocation, ultimately lowering stroke incidence and enhancing overall public health.

This section has described a full pipeline for stroke risk prediction from medical survey data, which includes data descriptions, preprocessing, feature engineering, model selection, training, validation, and evaluation detailed in **Fig 2**. A large survey dataset with an extensive number of feature columns was meticulously selected to predict stroke risk. Key predictive variables were carefully selected across seven distinctive data domains. The dataset, which was gathered from individual telephone responses, requires substantial preprocessing and comprehensive data analysis before it can be used in prediction models. Six different models, including both machine learning and deep learning paradigms, were thoroughly examined and the outcomes were compared. Based on the model performance evaluations, three standout models—soft voting, hard voting, and stacking—were chosen for model fusion [20] [21]. To evaluate the usefulness of these top-performing models, the performances were rigorously visualized using a variety of performance indicators, such as classification reports, confusion matrices, and AUC curves. Furthermore, Explainable AI (XAI) [22] [23] approaches are used to explicate the precise contributions of the selected characteristics within the dataset.

**Fig. 2:**
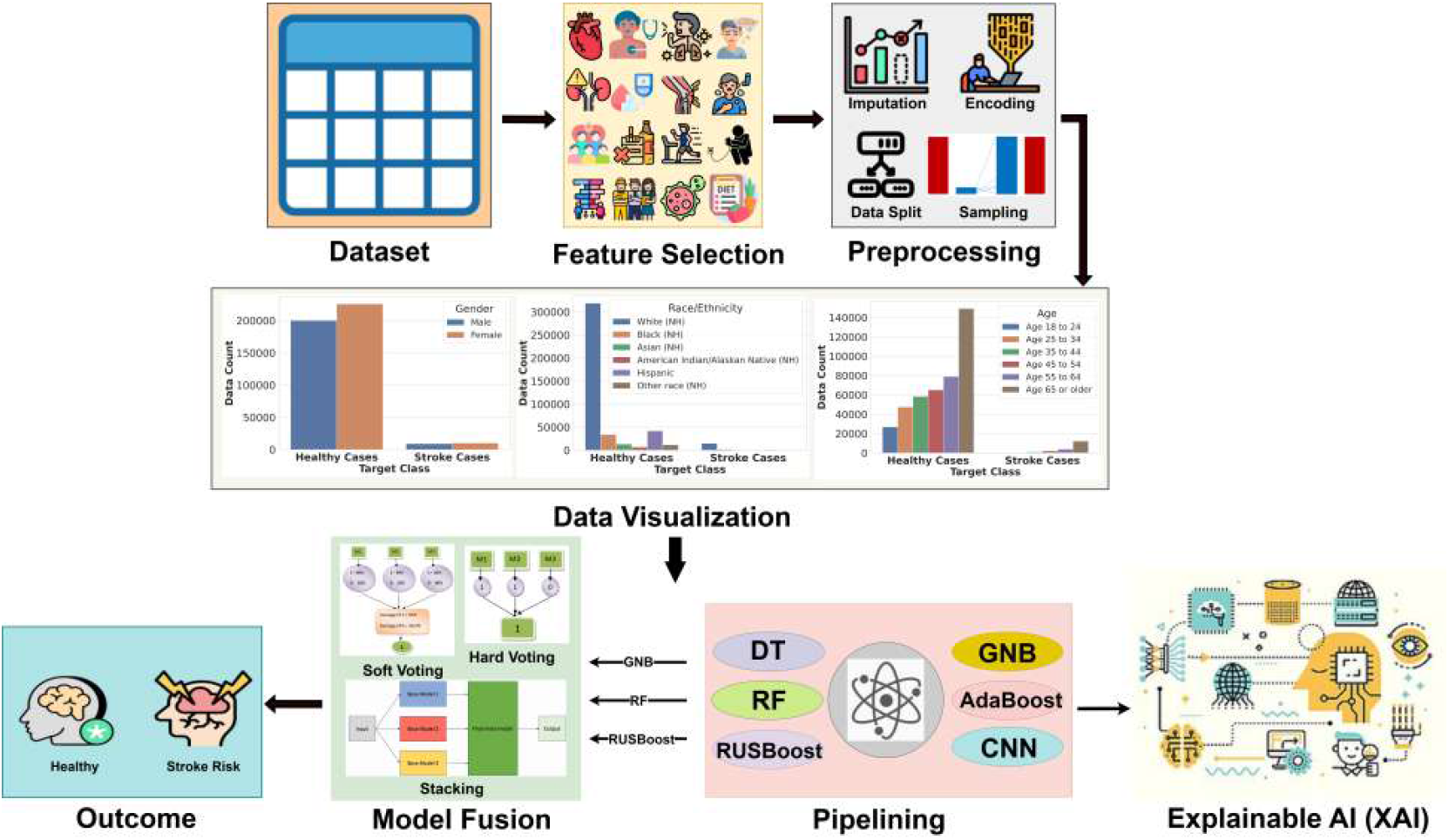
Representation of the complete workflow to address stroke risk prediction from medical survey data. All the required steps to acquire thorough analyses and comparisons of the proposed approaches in terms of stroke risk prediction are represented in this section.

## V. Feature selection

The relevance of feature selection in predicting the risk of stroke using BRFSS survey data cannot be overstated. Feature selection improves model performance by finding and maintaining the most important variables while removing redundant or unnecessary ones [16] [17]. This decreases computing complexity, allowing for more efficient and accurate risk evaluations.

The Behavioural Risk Factor Surveillance System (BRFSS) dataset contains data from several data domains and includes over 300 input characteristics in tota6 [11] [12] [16]. A thoughtful selection based on the prediction label (stroke risk prediction) was employed to include 40 essential input features drawn from seven different data domains specifically for the goal of conducting extensive analysis and prediction linked to strokes detailed in **Table I**. This thoughtful feature curation assures the analytical and predictive models’ applicability and performance in responding to stroke-related issues in the BRFSS dataset.

**TABLE I:** Illustration of the specified input features from several data domains. Selective features from seven input domains to perform stroke risk prediction have been presented in this section.

| Data Domain | Feature Description |
| --- | --- |
| Social and demographic factors | Marital status, Residential status, Military record, Ethnicity (Race), State, Spoken languages, Gender, Children count, Age group |
| Socio-economic status | Literacy status, Employment status, Earning level (Income), Mobile usage |
| Medical history | Skin cancer history, Other cancer history, Chronic bronchitis history, Depressive disorder history, Renal disease history (Kidney issues), Diabetes history, Arthritis history, Myocardial infarction history, Angina history, Asthma history, Self-rated Health status, BMI level |
| Disability status | Difficulty in seeing, Difficulty in dressing or bathing, Difficulty in walking, Difficulty in doing errands alone |
| Healthcare services | Health Insurance, Doctor affordability, Routine checkup, Availability of personal health care assistant |
| Personal health behavior | Alcohol consumption, Tobacco consumption, Smoking level, Exercise level |
| Vaccination history | HIV status, Vaccination records |

The SHapley Additive exPlanations (SHAP) [22] [23] approach is used in this section to uncover the inherent relevance of various characteristics within the selected dataset represented in **Fig 3**. The SHAP summary plot is used to visualize the significance by focusing exclusively on the top 24 significant variables. This graphical depiction gives useful insights for deciphering the complex interactions between these parameters and how these contribute to the prognosis of both healthy and Stroke patients. This study provides a thorough and intuitive knowledge of which specific features have the most effect in determining these particular outcomes, measuring the amount to which each feature contributes to the specific prediction it informs.

**Fig. 3:**
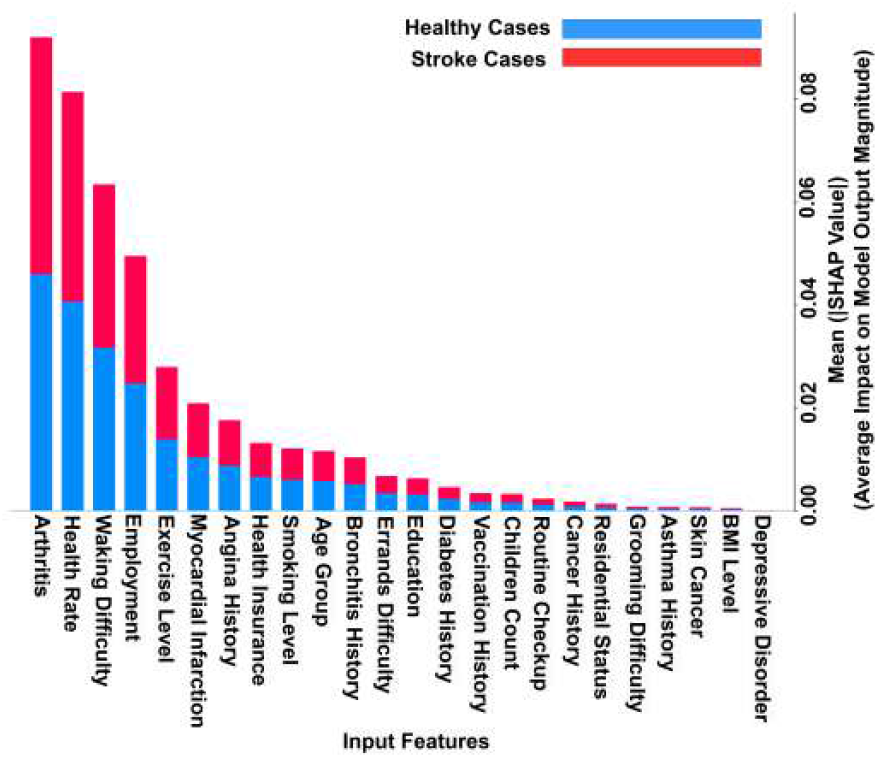
Illustration of deep insightful features importance using the SHapley Additive exPlanations (SHAP). The feature importance of the top 24 impactful features among the selected features is visualized in this section using the SHAP summary plot.

## VI. Preprocessing

When collecting survey data via telephone conversations, it is common to find that a significant fraction of the data is missing [11] [14]. To successfully resolve these missing data, proper imputation approaches for chosen characteristics must be employed. Accordingly, the target column requires encoding to aid modeling. The dataset is then required to be divided into separate training and testing sets for model assessment. Furthermore, sampling strategies could potentially used to address any observed class imbalance concerns in the dataset. These sequential stages are the necessary preprocessing operations that must be carried out prior to feeding the prepared data into the prediction models.

This section depicts the various preprocessing stages performed on the selected feature data to prepare it for input into prediction models detailed in **Fig 4**. The meticulously curated dataset is currently undergoing critical preprocessing stages to resolve missing values using Random Forest-based computations, which use existing insights to predict and impute feature-specific missing data. Label encoding is used to prepare the target column for prediction, with the target variable initially classified into four characteristics: input for healthy cases, input for stroke cases, refusal to provide responses, and individuals with no knowledge of the response. These variables are binary encoded, with stroke cases being the positive class and the rest indicating healthy instances. Following that, the data is divided into training and test sets to aid in the development and evaluation of prediction models. Notably, there is a class imbalance issue, with a much higher percentage of healthy instances compared to stroke cases. Synthetic Minority Over-sampling Technique (SMOTE) is used to balance the target classes within the training set to rectify this imbalance. The data has become prepared to be input into prediction models after these preparation processes have been performed.

**Fig. 4:**
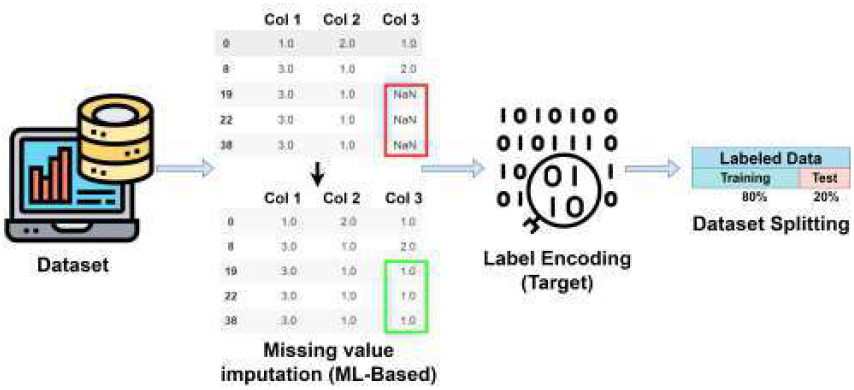
Illustration of the data preprocessing phases. The data with selected features require a number of preprocessing steps to be input into the prediction models. Hence, all the performed preprocessing steps are visualized in this section.

## VII. Experiment and result analysis

On an updated iteration of the Behavioural Risk Factor Surveillance System (BRFSS) dataset, a thorough study employing a wide combination of machine learning and deep learning approaches was carried out in the quest of optimizing stroke risk prediction [11] [14]. Traditional Decision Tree (DT) models, ensemble techniques like Random Under Sampling and Boosting (RusBoost), Random Forest (RF), Adaptive Boosting (AdaBoost), and Gaussian Nave Bayes (GNB), as well as sophisticated ensemble techniques like soft and hard Voting and Stacking with Logistic Regression (LR) as meta-classifier, were all included in the employed techniques. Incorporating a 1D Convolutional Neural Network (CNN) with residual networks, the study also delves into the field of deep learning. This diverse approach aims to methodically assess and compare the predictive performance of various models in relation to stroke risk prediction.

The effectiveness of different categorization procedures or models is evaluated in large part by performance matrices depicted in **Table II**. The train and test accuracies were presented pointing out the way proposed models are performed in terms of training and testing. However, it is crucial to emphasize that in such research, performance in predicting both the positive (stroke cases) and negative (healthy cases) classes is more important than high accuracy. The measured average values of precision, recall, and f1 score in this section provide a combined evaluation of the classification models’ overall performance, taking into consideration both the distinct class metrics and the respective class frequencies. Weighted averages are particularly significant as these offer a comprehensive assessment of a model’s capacity to accurately categorize samples across all classes. These metrics play a crucial role in assisting decision-makers in determining the best categorization technique choice for the particular task and enabling better informed and efficient decision-making processes.

**TABLE II:**
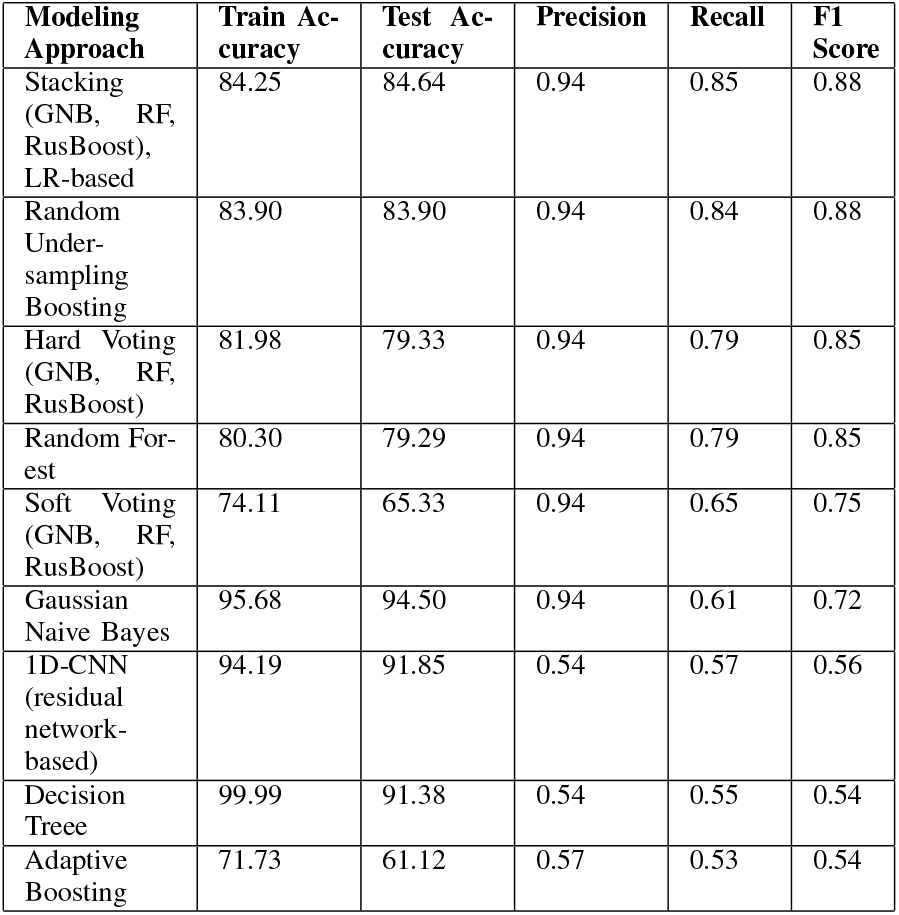
Illustration of classification report with train and test accuracy for all the proposed models. Calculated average values of precision, recall, and f1 score for all the proposed approaches along with acquired training and testing accuracies are presented in this section. The arrangement of the representation is in descending order of the F1 score.

| Modeling Approach | Train Accuracy | Test Accuracy | Precision | Recall | F1 Score |
| --- | --- | --- | --- | --- | --- |
| Stacking (GNB, RF, RusBoost), LR-based | 84.25 | 84.64 | 0.94 | 0.85 | 0.88 |
| Random Under-sampling Boosting | 83.90 | 83.90 | 0.94 | 0.84 | 0.88 |
| Hard Voting (GNB, RF, RusBoost) | 81.98 | 79.33 | 0.94 | 0.79 | 0.85 |
| Random Forest | 80.30 | 79.29 | 0.94 | 0.79 | 0.85 |
| Soft Voting (GNB, RF, RusBoost) | 74.11 | 65.33 | 0.94 | 0.65 | 0.75 |
| Gaussian Naive Bayes | 95.68 | 94.50 | 0.94 | 0.61 | 0.72 |
| 1D-CNN (residual network-based) | 94.19 | 91.85 | 0.54 | 0.57 | 0.56 |
| Decision Tree | 99.99 | 91.38 | 0.54 | 0.55 | 0.54 |
| Adaptive Boosting | 71.73 | 61.12 | 0.57 | 0.53 | 0.54 |

The formal formulation of the Area Under the Receiver Operating Characteristic (AUC) curve for specified models is presented in this section, demonstrating the significance of the AUC curve in measuring classifier performance detailed in **Fig 5**. The AUC curve depicts a classifier’s ability to differentiate between positive and negative occurrences across varied discriminating levels. Notably, the AUC measurements for the best-performing fusion models in the study outperform traditional classifiers such as Gaussian Naive Bayes, Random Forest, and RUSBoost. This highlights the fusion models’ better discriminative and prediction capability, indicating the potential to improve classification tasks across several domains.

**Fig. 5:**
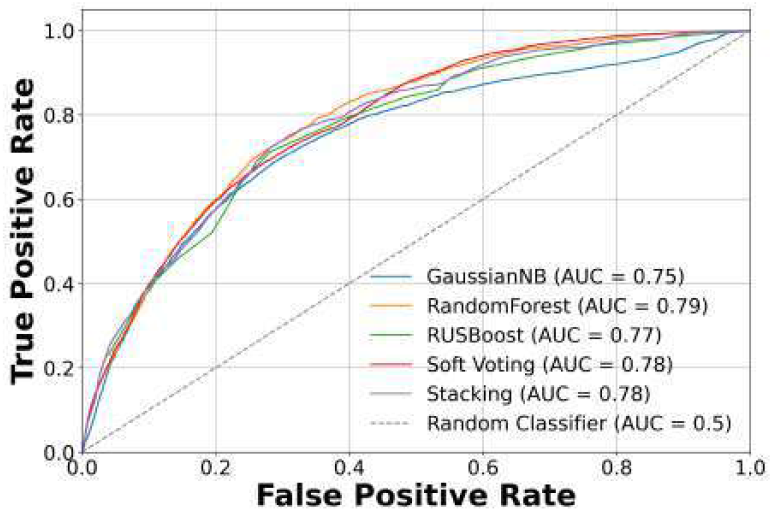
Representation of the Area Under the Receiver Operating Characteristic (AUC) curve for specified models. The AUC measures for the best-performing classifiers are represented in this section.

## VIII. Conclusion

Leveraging the quantity of data available in the CDC’s 2022 BRFSS dataset, this work marks a significant advancement in the field of stroke risk prediction. This has proved the predictive ability of the proposed models as well as the possibility for model fusion by methodically choosing important predictive variables across varied data domains. Furthermore, the application of Explainable AI (XAI) methodologies offered a layer of interpretability to acquired findings, revealing light on the distinct advantages of several characteristics of stroke risk prediction. Overall, the findings emphasize the requirement for early identification and treatment in individuals with stroke risk, with the ultimate objective of lowering stroke incidence and enhancing public health.

In conclusion, the suggested comprehensive strategy, which includes data preparation, model validation, and XAI-driven feature interpretation, establishes this work as a trailblazing effort in understanding stroke risk prediction. The findings of this study have the potential to inspire targeted strategies for awareness-raising, and healthcare resource allocation, ultimately assisting to reduce stroke-related disability and healthcare expenditures.

## Data Availability

The Behavioural Risk Factor Surveillance System (BRFSS)2022 dataset was used in this study,

https://www.cdc.gov/brfss/index.html

